# Reasoning Beyond Accuracy: Expert Evaluation of Large Language Models in Diagnostic Pathology

**DOI:** 10.1101/2025.04.11.25325686

**Authors:** Asim Waqas, Ehsan Ullah, Asma Khan, Farah Khalil, Zarifa Gahramanli Ozturk, Vaibhav Chumbalkar, Daryoush Saeed-Vafa, Zena Jameel, Weishen Chen, Humberto Trejo Bittar, Jasreman Dhillon, Rajendra S. Singh, Andrey Bychkov, Anil V. Parwani, Marilyn M. Bui, Matthew B. Schabath, Ghulam Rasool

**Affiliations:** Department of Cancer Epidemiology H. Lee Moffitt Cancer Center & Research Institute Tampa, FL; Department of Surgery Health New Zealand, Counties Manukau Auckland, New Zealand; Armed Forces Institute of Pathology Rawalpindi, Pakistan; Department of Pathology H. Lee Moffitt Cancer Center & Research Institute; Clinical Science Lab H. Lee Moffitt Cancer Center & Research Institute; Department of Dermatology & Cutaneous Surgery University of South Florida Tampa, FL; Department of Anatomic Pathology H. Lee Moffitt Cancer Center & Research Institute; Dermatopathology and Digital Pathology Summit Health, Berkley Heights, NJ; Department of Pathology Kameda Medical Center Kamogawa City, Chiba Prefecture, Japan; Department of Pathology The Ohio State University Columbus, Ohio; Department of Cancer Epidemiology H. Lee Moffitt Cancer Center & Research Institute; Department of Machine Learning H. Lee Moffitt Cancer Center & Research Institute

**Keywords:** Generative AI, Reasoning Large Language Models, Pathology, Clinical Reasoning, AI Evaluation

## Abstract

**Background:** Diagnostic pathology depends on complex, structured reasoning to interpret clinical, histologic, and molecular data. Replicating this cognitive process algorithmically remains a significant challenge. As large language models (LLMs) gain traction in medicine, it is critical to determine whether they have clinical utility by providing reasoning in highly specialized domains such as pathology.

**Methods:** We evaluated the performance of four reasoning LLMs (OpenAI o1, OpenAI o3-mini, Gemini 2.0 Flash Thinking Experimental, and DeepSeek-R1 671B) on 15 board-style open-ended pathology questions. Responses were independently reviewed by 11 pathologists using a structured framework that assessed language quality (accuracy, relevance, coherence, depth, and conciseness) and seven diagnostic reasoning strategies. Scores were normalized and aggregated for analysis. We also evaluated inter-observer agreement to assess scoring consistency. Model comparisons were conducted using one-way ANOVA and Tukey’s Honestly Significant Difference (HSD) test.

**Results:** Gemini and DeepSeek significantly outperformed OpenAI o1 and OpenAI o3-mini in overall reasoning quality (p < 0.05), particularly in analytical depth and coherence. While all models achieved comparable accuracy, only Gemini and DeepSeek consistently applied expert-like reasoning strategies, including algorithmic, inductive, and Bayesian approaches. Performance varied by reasoning type: models performed best in algorithmic and deductive reasoning and poorest in heuristic and pattern recognition. Inter-observer agreement was highest for Gemini (p < 0.05), indicating greater consistency and interpretability. Models with more in-depth reasoning (Gemini and DeepSeek) were generally less concise.

**Conclusion:** Advanced LLMs such as Gemini and DeepSeek can approximate aspects of expert-level diagnostic reasoning in pathology, particularly in algorithmic and structured approaches. However, limitations persist in contextual reasoning, heuristic decision-making, and consistency across questions. Addressing these gaps, along with trade-offs between depth and conciseness, will be essential for the safe and effective integration of AI tools into clinical pathology workflows.

## 1 Introduction

Anatomical pathologists draw upon years of training and accumulated clinical experience to develop a range of diagnostic reasoning strategies for interpreting histopathologic findings in the context of patient presentation. This interpretive process requires not only deep visual pattern recognition but also the ability to synthesize clinical, morphologic, and molecular data using a broad array of reasoning approaches [1]. These include algorithmic workflows, deductive and inductive (hypothetico-deductive) reasoning, Bayesian and heuristic inference, mechanistic understanding of disease processes, and pattern recognition—each contributing to the integrative synthesis of visual, clinical, and contextual cues [1].

With the advent of generative artificial intelligence (AI), large language models (LLMs) such as ChatGPT, Gemini, and DeepSeek have garnered interest for their potential to support pathology workflows [2, 3]. These models generate responses by leveraging probabilistic associations across vast biomedical corpora [4, 5]. Early applications in pathology have focused on clinical summarization [6], education, and simulation; however, most evaluations have assessed only the factual accuracy or superficial plausibility of outputs, without examining the structure or soundness of the underlying reasoning processes [7, 8, 9, 10].

Although LLMs have shown strong performance on general medical benchmarks, their ability to replicate the nuanced, domain-specific reasoning required in diagnostic pathology remains unclear [11, 12, 13]. Pathology demands more than knowledge retrieval—it requires the contextual application of varied reasoning strategies across morphologic, molecular, and clinical dimensions. Given that board licensing examinations emphasize reasoning beyond factual recall [14], a critical question emerges: can LLMs generate responses that are not only accurate but also reflect expert-like diagnostic reasoning?

To address this question, we developed a novel, structured evaluation framework grounded in clinical practice. Eleven expert pathologists evaluated responses generated by four LLMs to 15 open-ended diagnostic questions. Each response was scored across five language quality metrics and seven clinically relevant reasoning strategies [1, 14]. The models, OpenAI o1, OpenAI o3-mini, Gemini 2.0 Flash Thinking Experimental (Gemini), and DeepSeek-R1 671B (DeepSeek), were compared across all evaluation criteria. Inter-observer agreement analysis was conducted to assess consistency among expert reviewers. Together, these findings offer a comprehensive assessment of clinical reasoning in LLMs and establish a scalable, domain-specific framework for evaluating AI systems in diagnostic pathology.

## 2 Materials and Methods

In this study, a structured evaluation framework was developed to assess the capacity of LLMs to generate clinically relevant, well-reasoned answers to open-ended pathology questions. Our focus was on both language quality and diagnostic reasoning, with expert review by pathologists. Based on the study by Wang et al. [14], a set of 15 open-ended diagnostic questions, listed in Supplementry Table 1, was selected to reflect the complexity and format of board licensing examinations.

### 2.1 Model Selection and Response Generation

Four state-of-the-art LLMs were evaluated: OpenAI o1, OpenAI o3-mini, Gemini, and DeepSeek. Each model was prompted independently to respond to all 15 questions in a zero-shot setting. To simulate a high-stakes clinical reasoning task, each question was preceded by the instruction: “This is a pathology-related question at the level of licensing (board) examinations”. No additional context or few-shot examples were provided. Model responses were collected between February and March 2025 using publicly accessible web interfaces or Application Programming Interface (API) endpoints. Each output included both a direct answer and explanatory reasoning. No fine-tuning or post-processing was applied to model responses.

### 2.2 Evaluation by Pathologists

Eleven expert pathologists (qualifications provided in Supplementary Table 2) independently evaluated the reasoning outputs generated by each LLM across 15 diagnostic questions. To mitigate potential bias, all evaluations were conducted in a blinded manner, and inter-observer agreement was analyzed to ensure consistency among evaluators. The evaluation assessed two complementary dimensions: the linguistic quality of the responses and the application of pathology-specific diagnostic reasoning strategies. The evaluation focused on the following two primary criteria:

1. Language Quality Metrics: Assessments were conducted on the clarity, coherence, depth, accuracy, and conciseness of the responses. These metrics are commonly employed in evaluating LLM outputs to ensure that generated content is not only factually correct but also well-structured and comprehensible. Such evaluation frameworks have been discussed in studies analyzing LLM performance in medical contexts [15].
2. Application of Pathology-Specific Reasoning Strategies: Evaluators examined the extent to which LLMs appropriately utilized various diagnostic reasoning approaches integral to pathology practice. These approaches include pattern recognition, algorithmic reasoning, inductive hypothetico-deductive reasoning, mechanistic insights, deductive reasoning, heuristic reasoning, and probabilistic (Bayesian) reasoning (details provided in Supplemental Table 3). The framework for these reasoning strategies is grounded in the cognitive processes outlined by Pena and Andrade-Filho, who describe the diagnostic process as encompassing cognitive, communicative, normative, and medical conduct domains [1, 16].

### 2.3 Statistical Analysis

Scores (on a 1–5 Likert scale) were first normalized to a 0–1 range using linear scaling (score divided by 5). Following normalization, average scores were computed across raters for each combination of model, question, and evaluation criterion. These aggregated values were used to evaluate model-level and criterion-specific performance. One-way analysis of variance (ANOVA) was used to assess differences in mean scores across models for each evaluation metric, including cumulative scores. When statistically significant, pairwise comparisons were conducted using Tukey’s Honestly Significant Difference (HSD) test (*α* = 0.05). Analyses were conducted using Python (v3.11) with standard statistical libraries. To assess inter-observer reliability, the percent agreement was calculated for each Question–Model–Criterion (Q-M-C) combination, which was defined as the proportion of raters who selected the most common score. Ratings ranged from 7 to 11 per Q-M-C (mean = 9.9). Model-level differences in percent agreement were tested using the Kruskal–Wallis H test. When significant, post hoc pairwise comparisons were performed using Dunn’s test with Bonferroni correction for multiple testing. Additional analyses, including normalization procedures, missing data summaries, and full pairwise comparison results, are available in Supplementary Tables 4, 5, 6, 7, 8, and 9 and Figures 6 and 7.

## 3 Results

We evaluated the performance of four LLMs (Gemini, DeepSeek, OpenAI o1, and OpenAI o3-mini) on 15 expert-generated diagnostic pathology questions using our structured evaluation framework. Responses were assessed across two primary domains: *Language Quality and Response Structure*, focused on NLP-oriented metrics (e.g., relevance, coherence, accuracy); and *Diagnostic Reasoning Strategies*, capturing clinically grounded reasoning styles (e.g., pattern recognition, algorithmic reasoning, mechanistic insight). Each domain includes a cumulative score and multiple sub-metrics, yielding twelve evaluation criteria overall.

Fig. 2 summarizes model performance across these criteria using radar plots. Gemini clearly outperforms all other models on language quality metrics (Fig. 2a), with the largest margins in “depth” and “coherence”. DeepSeek ranks second overall, while the two OpenAI reasoning models show similar performance, particularly on accuracy and relevance. Fig. 2b visualizes pathology-oriented reasoning strategies. Gemini again leads across most dimensions, especially algorithmic and inductive reasoning. DeepSeek demonstrates competitive performance, whereas the OpenAI models score lower in mechanistic, heuristic, and Bayesian reasoning, suggesting less robust clinical reasoning capabilities. These overarching trends are explored in greater detail in the subsequent sections.

**Figure 1:**
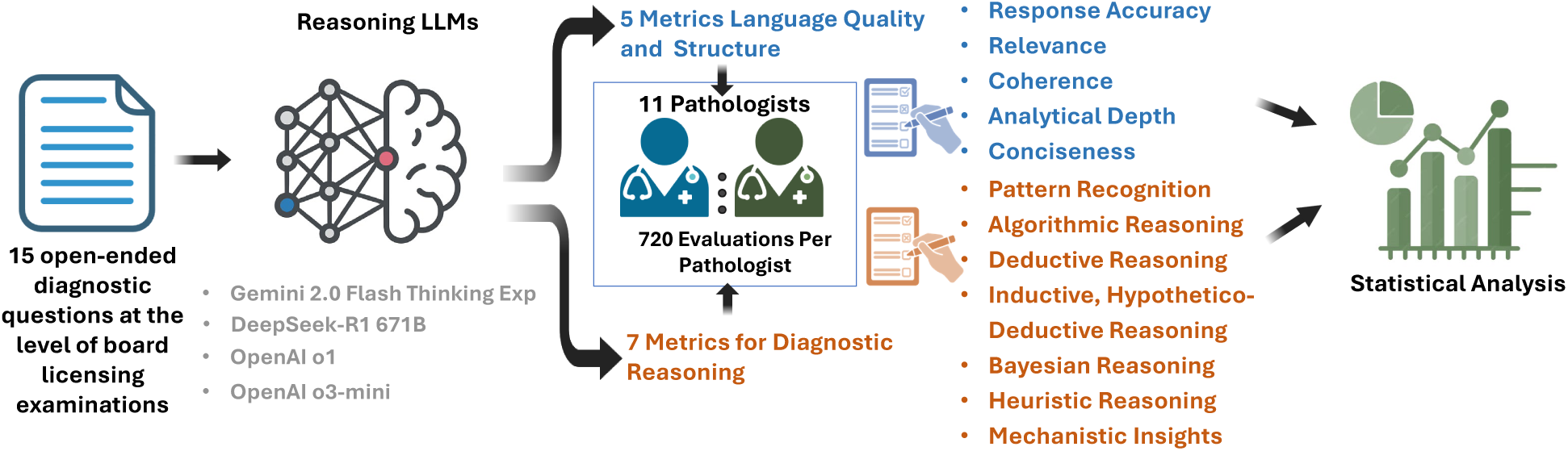
Evaluation Framework for Assessing Diagnostic Reasoning in Large Language Models. Fifteen open-ended diagnostic pathology questions, reflecting the complexity of board licensing examinations, were independently submitted to four LLMs: OpenAI o1, OpenAI o3-mini, Gemini 2.0 Flash-Thinking Experimental (Gemini), and DeepSeek-R1 671B (DeepSeek). Each response was evaluated by 11 expert pathologists using a structured rubric comprising 12 metrics across two domains: (1) language quality and response structure (relevance, coherence, accuracy, depth, and conciseness) and (2) diagnostic reasoning strategies (pattern recognition, algorithmic, deductive, inductive/hypothetico-deductive, heuristic, mechanistic, and Bayesian reasoning). Pathologists were blinded to model identification. Evaluation scores were aggregated and normalized to account for missing data and served as the basis for both model performance comparisons and inter-observer agreement analysis.

**Figure 2:**
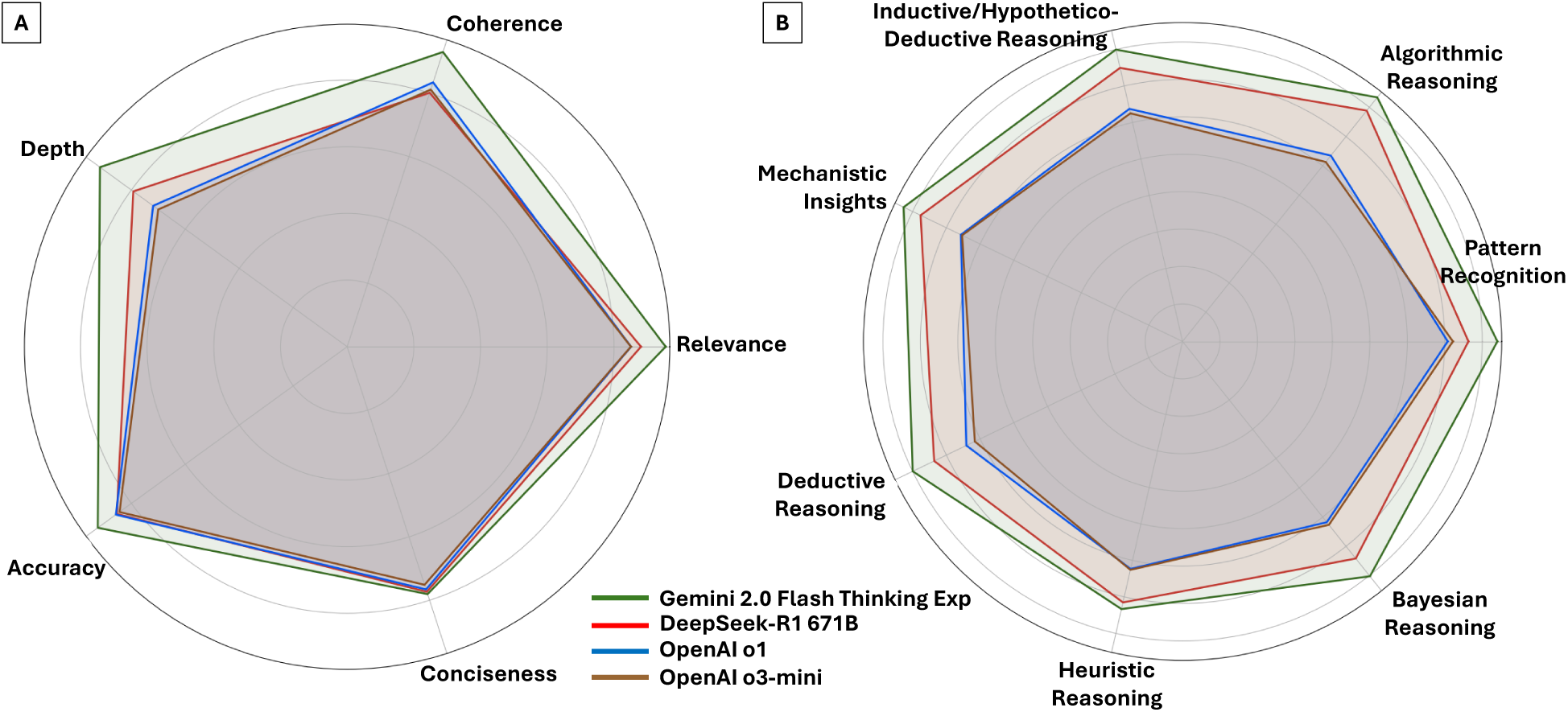
Comparative Performance of LLMs Across Language Quality and Diagnostic Reasoning Domains. Radar plots summarize the normalized mean scores (range: 0 to 1) assigned by 11 pathologists for each model across 12 evaluation criteria. **Panel A**: Language quality metrics, including accuracy, relevance, analytical depth, coherence, conciseness, and cumulative scores. **Panel B**: Diagnostic reasoning strategies, including pattern recognition, algorithmic reasoning, deductive reasoning, inductive/hypothetico-deductive reasoning, heuristic reasoning, mechanistic insights, and Bayesian reasoning. Gemini consistently outperformed other models across both domains, particularly in analytical depth and structured reasoning. OpenAI o1 showed the greatest variability in performance across metrics.

### 3.1 Evaluation of Language Quality and Response Structure

Fig. 3 presents normalized model performance across five language-focused evaluation metrics—relevance, coherence, analytical depth, accuracy, and conciseness—along with a cumulative average score. Fig. 3A shows cumulative scores across all 15 questions, while Figs. 3B–F display model-specific performance on individual metrics. Across all dimensions, Gemini consistently achieved the highest average scores. In cumulative performance (Fig. 3A), Gemini significantly outperformed OpenAI o1, OpenAI o3-mini, and DeepSeek (p < 0.05). DeepSeek also significantly outperformed both OpenAI models (p < 0.05), while there was no significant difference between OpenAI o1 and OpenAI o3-mini (p > 0.05).

**Figure 3:**
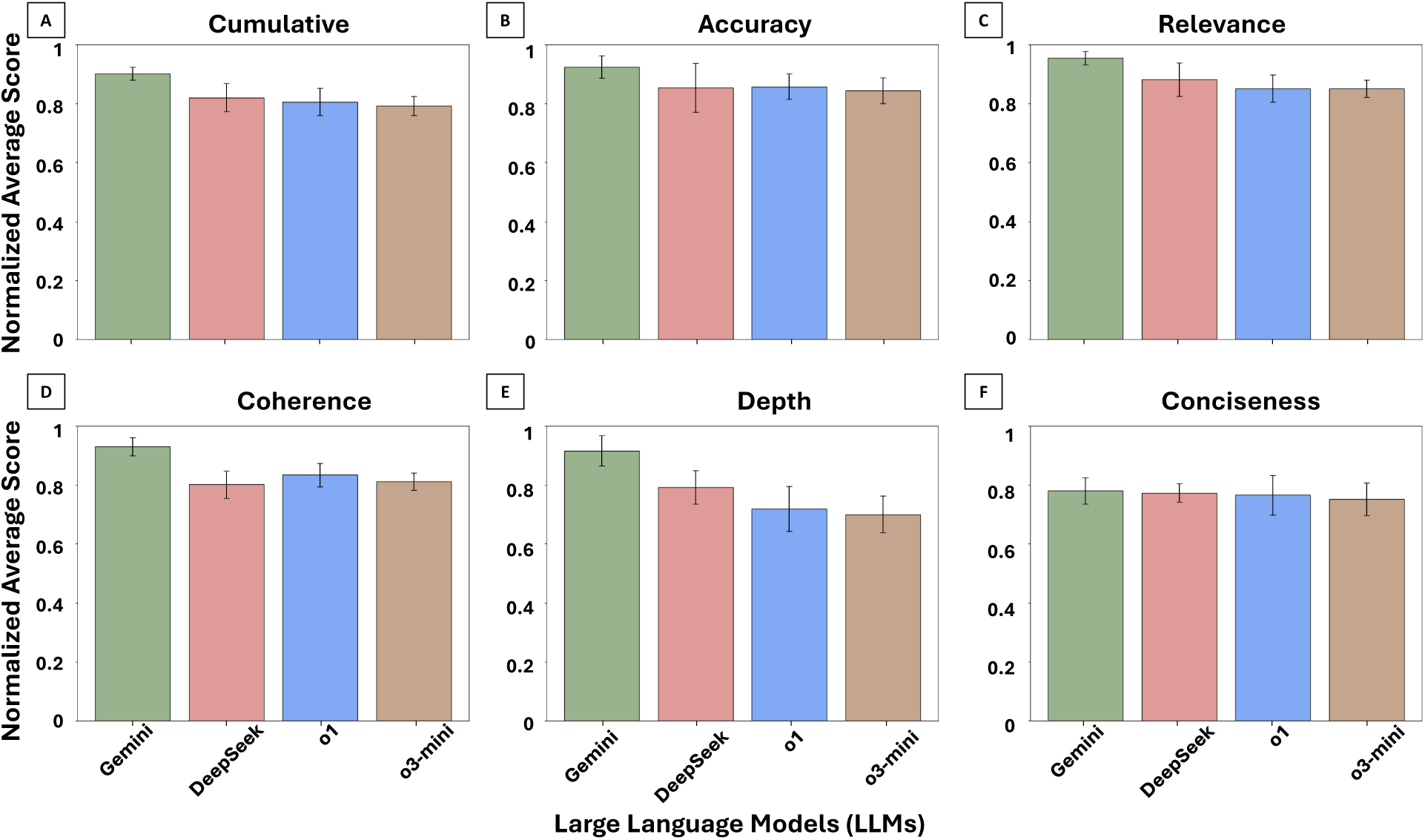
LLM Performance on Language Quality Metrics. Normalized average scores (range: 0 to 1) across five core language quality dimensions—accuracy, relevance, coherence, analytical depth, and conciseness—based on ratings from 11 pathologists across 15 pathology questions. **Panels A–F**: Overall average performance (A), followed by Accuracy (B), Relevance (C), Coherence (D), Analytical Depth (E), and Conciseness (F). Gemini consistently achieved the highest scores across all metrics, with the greatest variability observed in coherence and analytical depth.

For accuracy (Fig. 3B), Gemini significantly outperformed both DeepSeek and OpenAI o1 (p < 0.05). All other pairwise differences were not statistically significant (p > 0.05). In relevance (Fig. 3C), all models maintained relatively high scores, with Gemini significantly outperforming all others (p < 0.05). No significant differences were observed among DeepSeek, OpenAI o1, and OpenAI o3-mini (p > 0.05). For coherence (Fig. 3D), Gemini significantly outperformed all three other models (p < 0.05), while no statistically significant differences were observed between DeepSeek and either OpenAI model (p > 0.05).

Analytical depth (Fig. 3E) showed more variation; Gemini significantly outperformed all other models (p < 0.05), and DeepSeek significantly outperformed both OpenAI o1 and OpenAI o3-mini (p < 0.05). No significant difference was observed between the two OpenAI models (p > 0.05). Lastly, conciseness (Fig. 3F) showed comparable performance across all models. Although Gemini had slightly higher average scores, no statistically significant differences were observed across any pairwise comparisons (p > 0.05). Overall, these results suggest that LLMs varied most on metrics requiring deeper interpretive judgment (e.g., analytical depth and coherence), with more consistent performance observed on factual dimensions like relevance and accuracy.

### 3.2 Evaluation of Diagnostic Reasoning Strategies

Fig. 4 shows model performance across seven clinically relevant diagnostic reasoning strategies: pattern recognition, algorithmic reasoning, deductive reasoning, inductive/hypothetico-deductive reasoning, Bayesian reasoning, heuristic reasoning, and mechanistic insights. Fig. 4A presents cumulative reasoning scores averaged across all strategies and questions, while Fig. 4B–H display model-specific performance on each individual reasoning type. In overall performance (Fig. 4A), Gemini achieved the highest cumulative score, followed by DeepSeek, with both OpenAI models scoring lower. Pairwise comparisons showed statistically significant differences between Gemini and all other models (p < 0.05), as well as between DeepSeek and both OpenAI variants (p < 0.05). No significant difference was found between OpenAI o1 and OpenAI o3-mini (p > 0.05).

**Figure 4:**
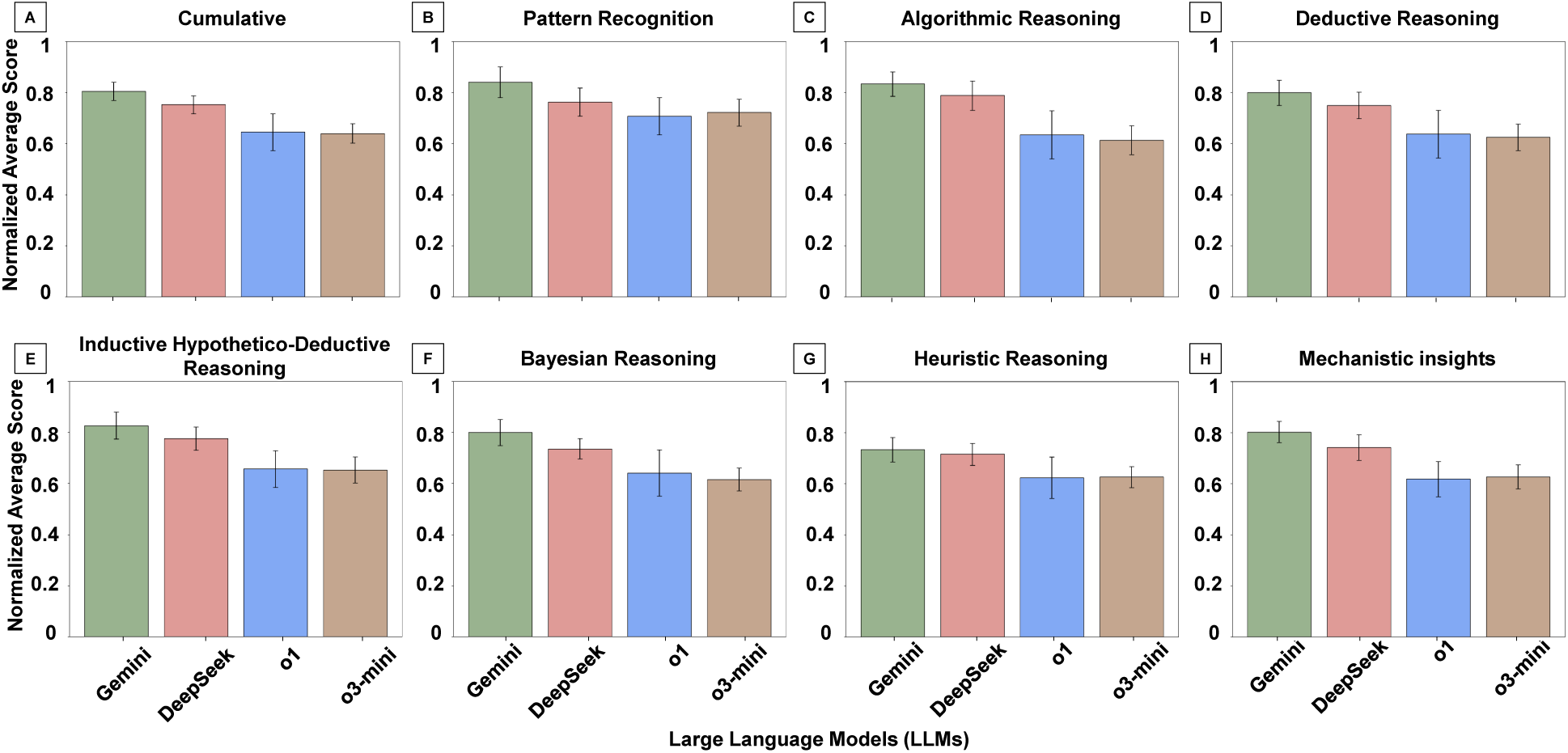
Diagnostic Reasoning Performance by Reasoning Type. Normalized mean scores (range: 0 to 1) for each LLM across seven diagnostic reasoning strategies based on expert evaluation of pathology-related questions. **Panels A–H**: Cumulative reasoning performance (A), Pattern Recognition (B), Algorithmic Reasoning (C), Deductive Reasoning (D), Inductive/Hypothetico-Deductive Reasoning (E), Bayesian Reasoning (F), Heuristic Reasoning (G), and Mechanistic Insights (H). Gemini and DeepSeek consistently outperformed the OpenAI models across most reasoning types, with particularly strong performance in algorithmic, inductive, and mechanistic reasoning. Heuristic and Bayesian reasoning yielded the lowest scores across all models, reflecting challenges with uncertainty-driven and experiential inference.

In pattern recognition (Fig. 4B), Gemini significantly outperformed all other models (p < 0.05), while differences between DeepSeek and the OpenAI models were not statistically significant (p > 0.05). For algorithmic reasoning (Fig. 4C), which yielded the highest scores among all reasoning types, Gemini and DeepSeek both significantly outperformed OpenAI o1 and OpenAI o3-mini (p < 0.05), though no difference was observed between Gemini and DeepSeek (p > 0.05). In deductive reasoning (Fig. 4D), Gemini significantly outperformed both OpenAI models (p < 0.05), and DeepSeek also significantly outperformed both OpenAI o1 and OpenAI o3-mini (p < 0.05). No significant difference was observed between Gemini and DeepSeek or between the two OpenAI models. For inductive/hypothetico-deductive reasoning (Fig. 4E), both Gemini and DeepSeek significantly outperformed the OpenAI models (p < 0.05), with no difference observed between Gemini and DeepSeek (p > 0.05).

Bayesian (probabilistic) reasoning scores (Fig. 4F) were generally lower across all models. However, all pairwise comparisons were statistically significant (p < 0.05), with Gemini outperforming all others and DeepSeek significantly outperforming both OpenAI models. In heuristic reasoning (Fig. 4G), Gemini and DeepSeek significantly outperformed both OpenAI models (p < 0.05) but did not significantly differ from each other (p > 0.05). No significant difference was observed between OpenAI o1 and o3-mini (p > 0.05). In mechanistic insights (Fig. 4H), all pairwise comparisons were statistically significant (p < 0.05). Gemini significantly outperformed DeepSeek and both OpenAI models, while DeepSeek significantly outperformed both OpenAI variants.

### 3.3 Inter-observer Percent Agreement Across Models

Percent agreement was analyzed for 720 unique Q-M-C combinations across four models: Gemini, DeepSeek, OpenAI o1, and OpenAI o3-mini. The overall mean percent agreement was 0.49 (range: 0.25-0.91). Fig. 5A shows a full distribution of percent agreement, with model-specific distributions shown in Fig. 5B. Percent agreement significantly differed across models (Kruskal-Wallis H = 76.798, p < 0.001). Post-hoc pairwise Dunn’s tests revealed that Gemini had significantly higher percent agreement than all other models (p < 0.001 for all comparisons). No significant differences were found among DeepSeek, OpenAI o1, and OpenAI o3-mini (p > 0.05).

**Figure 5:**
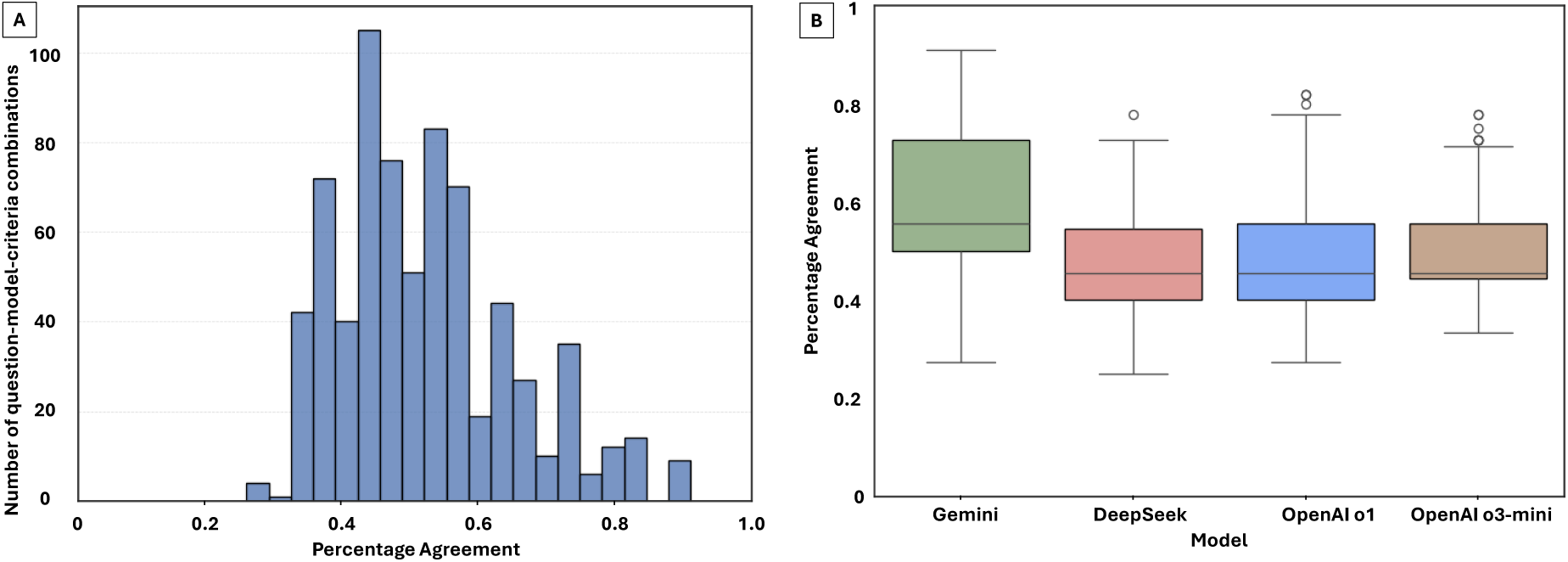
Percent agreement across 720 unique combinations of question, model, and evaluation criterion (Q–M–C), reflecting the proportion of raters who selected the most common score. **Panel A**: Distribution of percent agreement across all Q–M–C combinations. **Panel B**: Model-specific distributions of agreement. Gemini achieved significantly higher inter-observer agreement compared to all other models (p < 0.001), suggesting greater consistency and interpretability of its outputs. No statistically significant differences were observed in pair-wise testing between DeepSeek, OpenAI o1, and OpenAI o3-mini.

## 4 Discussion

This study evaluated the ability of four state-of-the-art LLMs to perform pathology-specific diagnostic reasoning using expert assessments of both language quality and clinical reasoning strategies. While all models produced generally relevant and accurate responses, only Gemini and DeepSeek demonstrated consistent strength across multiple reasoning dimensions, particularly analytical depth, coherence, and algorithmic reasoning. Moreover, Gemini achieved the highest inter-observer agreement among expert pathologists, suggesting greater clarity and interpretability in its responses. These findings highlight the need to move beyond accuracy-focused evaluation and toward models that generate clinically coherent and contextually intelligible reasoning, especially in high-stakes diagnostic domains such as pathology. This analysis also revealed substantial variation in the reasoning performance of LLMs when applied to pathology-focused diagnostic questions. Among the four evaluated models, Gemini consistently achieved the highest scores across both evaluation domains (language quality and diagnostic reasoning). DeepSeek followed closely in most categories, while the OpenAI variants (o1 and o3-mini) generally performed lower compared to the other LLMs, particularly in analytical depth, coherence, and advanced reasoning types.

In the language quality domain, all models produced broadly relevant responses. Gemini and DeepSeek distinguished themselves in analytical depth, coherence, and cumulative performance. Gemini also significantly outperformed other models in accuracy, suggesting more robust and precise responses. While the OpenAI models tended to be more concise, these differences were not statistically significant and often came at the expense of analytical depth and interpretability. In the diagnostic reasoning domain, all models performed best on algorithmic and deductive reasoning tasks. Gemini consistently outperformed all others across most reasoning types. DeepSeek also performed well in several categories, though its advantage over the OpenAI models was less pronounced in pattern recognition. Performance dropped markedly for all models in nuanced reasoning types such as heuristic, mechanistic, and Bayesian reasoning. These areas typically require integrating implicit knowledge, clinical judgment, and experience-based decision-making.

Notably, inter-observer agreement was highest for Gemini, indicating that its responses were more consistently interpreted and rated by expert pathologists. This suggests that Gemini’s outputs may be not only more complete or coherent but also more reliably understandable to clinical end-users. Such consistency is essential for downstream applications of LLMs in real-world diagnostic settings, where human-AI collaboration depends on shared reasoning clarity.

The prior evaluations of LLMs in medicine have focused on factual correctness, clinical acceptability, or performance on standardized assessments such as the USMLE and other multiple-choice examinations [11, 12, 13, 17]. While these studies demonstrate that advanced models can retrieve and generate clinically accurate information, they typically frame evaluation in binary terms, right or wrong, without assessing the structure, context, or interpretability of the underlying reasoning processes. However, recent studies have begun to explore how generative AI might support medical reasoning in pathology and related domains. Waqas et al. have outlined the promise of LLMs and FMs for digital pathology applications such as structured reporting, clinical summarization, and educational simulations [8, 9]. Brodsky et al. emphasized the importance of interpretability and reasoning traceability in anatomic pathology, noting that AI outputs must not only be accurate but also reflect logical, clinically meaningful reasoning patterns [7]. However, these studies primarily describe use cases or propose conceptual frameworks rather than empirically evaluating whether LLMs demonstrate structured reasoning aligned with human diagnostic strategies. Few studies have assessed whether model outputs reflect the types of reasoning, such as algorithmic, heuristic, or mechanistic, that are integral to expert pathology practice.

Histologic diagnosis is a cognitively demanding task that requires the integration of diverse information sources, ranging from visual features on tissue slides to clinical history and disease biology. As part of this decision-making process, pathologists rely on a broad set of reasoning strategies, including pattern recognition, algorithmic workflows, hypothetico-deductive reasoning, Bayesian inference, and heuristics, often applied fluidly based on experience and context [1, 18]. Despite their central role in diagnostic workflows, these reasoning approaches are rarely formalized in LLM evaluation studies. While prior cognitive and educational research has documented these strategies in pathology and clinical decision-making [19, 20, 21], few studies have translated them into a structured framework for evaluating AI systems in this domain. This study addresses this gap by developing and applying a structured evaluation framework that operationalizes seven clinically grounded reasoning strategies, i.e., pattern recognition, algorithmic reasoning, deductive and inductive reasoning, Bayesian reasoning, mechanistic insights, and heuristic reasoning. In contrast to prior work that has focused on factual accuracy or subjective plausibility, we used expert raters to assess whether model responses actually reflected reasoning types that pathologists would apply during their decision-making process. We also included five language quality metrics to assess the structure and interpretability of responses, features that are critical for clinical adoption but often overlooked in model evaluation. This approach builds on prior conceptual work in generative AI for pathology [7, 8, 9], but moves beyond descriptive analysis to provide a domain-specific, empirically tested benchmark for reasoning assessment.

The results of this study show that LLMs perform relatively well in algorithmic and deductive reasoning, likely reflecting their ability to retrieve structured clinical knowledge from training data and guidelines [12, 22, 23, 24]. However, model performance was markedly lower in reasoning types that rely more heavily on experience, context, and uncertainty, such as heuristic reasoning, Bayesian inference, and mechanistic understanding [25, 26]. These findings align with previous observations that current LLMs struggle with nuanced clinical reasoning, particularly in settings that require adaptive judgment [27, 28, 29, 30]. Moreover, our analysis highlights the lack of metacognitive regulation in LLMs, their inability to recognize uncertainty, self-correct, or reconcile conflicting information, challenges that have been noted in other domains as well [31]. By incorporating expert adjudication, reasoning taxonomies, and inter-observer agreement, our study offers a new model for evaluating clinical reasoning in generative AI that extends beyond correctness to assess the fidelity and structure of reasoning itself.

It is also important to interpret lower scores in certain reasoning categories in the context of clinical relevance and question content. For example, pattern recognition may be inapplicable to questions focused solely on molecular alterations or classification schemes, where visual cues are absent. Similarly, while heuristic reasoning was consistently rated low across models, some pathologists noted that this may reflect an appropriate aversion to premature conclusions based on past experience alone, an approach discouraged in modern diagnostic workflows that emphasize comprehensive differential diagnosis. These nuances underscore the importance of context when evaluating reasoning strategies.

Our findings have important implications for the safe and effective integration of LLMs into clinical workflows, particularly in diagnostic fields such as pathology, where interpretability, trust, and domain-specific reasoning are paramount. While all four models evaluated in this study produced largely relevant and accurate responses, only Gemini, and to a slightly lesser extent DeepSeek, demonstrated reasoning structures that aligned with real-world diagnostic approaches. These models also achieved higher inter-observer agreement among expert pathologists, suggesting that their outputs were not only well-structured but also consistently interpretable across users. This consistency is critical in clinical environments where diagnostic decisions are collaborative and where AI-generated insights must be readily understood, validated, and acted upon by human experts. In contrast, models with lower coherence and reasoning clarity may increase cognitive burden, introduce ambiguity, or erode trust, especially when deployed in high-stakes or time-constrained settings. By highlighting variation in reasoning quality across models and across different types of diagnostic logic, our results suggest that LLM evaluation should go beyond factual correctness and consider reasoning fidelity as a core requirement. This is particularly relevant for educational, triage, or decision support applications in pathology, where nuanced reasoning is essential, and automation must complement, rather than obscure, human expertise.

We acknowledge that this study has some limitations. Although a diverse set of reasoning strategies was evaluated, this analysis was limited to 15 open-ended questions. While these were curated by expert pathologists, they may not fully capture the range of diagnostic challenges encountered in practice. Future iterations of this benchmark should include a broader array of cases, including rare and diagnostically complex scenarios. We focused solely on text-based LLMs and did not assess multimodal models that incorporate pathology images. The use of expert raters (i.e., pathologists) adds clinical realism but also introduces subjectivity. We also did not measure downstream clinical impact, such as improvements in diagnostic accuracy or workflow efficiency. Future studies should explore these outcomes prospectively. In addition, the effects of prompting strategies, fine-tuning, and domain adaptation on reasoning performance deserve further investigation. Reasoning-aligned LLMs may hold promise for educational use, simulation, and decision support, applications that require careful validation in real-world settings.

## 5 Conclusion

This study presents a structured evaluation of advanced reasoning LLMs in pathology, focusing not only on factual accuracy but also on diagnostic reasoning quality. By assessing model outputs across five language metrics and seven clinically grounded reasoning strategies, we provide a nuanced benchmark that goes beyond traditional correctness-based evaluations. These findings demonstrate that while all models produce largely relevant and accurate responses, substantial differences exist in their ability to emulate expert diagnostic reasoning. Gemini and DeepSeek outperformed other models in both reasoning fidelity and inter-observer agreement, suggesting greater alignment with clinical expectations. Conversely, current limitations in heuristic, mechanistic, and probabilistic reasoning highlight areas for model improvement. To our knowledge, this is the first study to systematically evaluate the diagnostic reasoning capabilities of LLMs in pathology using expert-annotated criteria grounded in clinical practice. As generative AI tools continue to evolve, clinically meaningful evaluation frameworks, such as the one introduced here, will be essential for guiding safe and effective integration into diagnostic practice. This work lays the foundation for future studies that seek to assess not just whether models are right but whether they reason like clinicians.

## Data Availability

All data produced in the present study are available upon request to the authors.

## Acknowledgments

The research was supported in part by a Cancer Center Support Grant at the H. Lee Moffitt Cancer Center & Research Institute, awarded by the NIH/NCI (P30-CA76292), a Florida Biomedical Research Grant (21B12), an NIH/NCI grant (U01-CA200464), NSF Awards 2234836 and 2234468 and NAIRR pilot funding.

## Supplementary 1: Details of Evaluated Large Language Models

### Gemini 2.0 Flash Thinking Experimental

Gemini 2.0 Flash Thinking Experimental is an advanced AI model developed by Google DeepMind. This model is designed to balance reasoning capabilities with speed by employing an internal "thinking process" during response generation. This approach enhances the model’s ability to handle complex tasks, particularly in mathematics, science, and multimodal reasoning. Benchmark evaluations have demonstrated significant improvements over previous models, showcasing enhanced performance and explainability. Developers can access this experimental model through the Gemini API in Google AI Studio. Key features include: (1) Enhanced Reasoning: Utilizes an internal "thinking process" to improve problem-solving capabilities. (2) Speed and Efficiency: Optimized to balance complex reasoning with rapid response generation. (3) Multimodal Capabilities: Excels in tasks involving text, code, and images. Details are available at: https://deepmind.google/technologies/gemini/flash-thinking/

### DeepSeek-R1 671B

DeepSeek-R1 671B is an LLM developed by the Chinese AI startup DeepSeek. Released in January 2025, it features a Mixture-of-Experts (MoE) architecture with a total of 671 billion parameters, of which 37 billion are activated per token during inference. This design enhances resource efficiency without compromising performance. DeepSeek-R1 has demonstrated capabilities comparable to leading models in tasks such as mathematics, coding, and complex reasoning. Key features include: (1) Mixture-of-Experts Architecture: Efficiently utilizes a subset of parameters during inference for optimized performance. (2) High Parameter Count: Among the largest open-source LLMs, facilitating advanced reasoning tasks. (3) Open-Source Availability: Supports the research community with accessible model weights and code. Further details are available at: https://huggingface.co/deepseek-ai/DeepSeek-R1

### OpenAI o1

OpenAI o1 is a reasoning-focused AI model developed by OpenAI, officially released on December 5, 2024. It is designed to allocate additional processing time before generating responses, thereby improving performance on complex tasks, including science, coding, and mathematics. The model supports chain-of-thought prompting and multi-step reasoning, enhancing its interpretability and accuracy. Key features include: (1) Deliberative Processing: Spends more time "thinking" before responding to enhance reasoning quality. (2) Chain-of-Thought Prompting: Capable of breaking down complex problems into intermediate steps. (3) Versatile Applications: Excels in tasks requiring deep understanding and logical analysis. Further details are available at: https://openai.com/o1/

### OpenAI o3-mini

OpenAI o3-mini is a compact reasoning model introduced by OpenAI on January 31, 2025. It aims to provide enhanced reasoning capabilities with reduced computational requirements, making it suitable for applications where resources are limited. Despite its smaller size, o3-mini can outperform o1 in coding and other reasoning tasks, offering a balance between performance and efficiency. Key features include: (1) Resource Efficiency: Optimized for lower computational overhead without significant performance trade-offs. (2) Adjustable Reasoning Effort: Offers settings to balance speed and depth of reasoning. (3) Specialized Domains: Particularly adept in STEM-related tasks, including coding and mathematics. Further details are available: https://openai.com/index/openai-o3-mini/

## Supplementary 2. Pathology Board-Style Questions and Qualifications of Pathologists

**Table 1:**
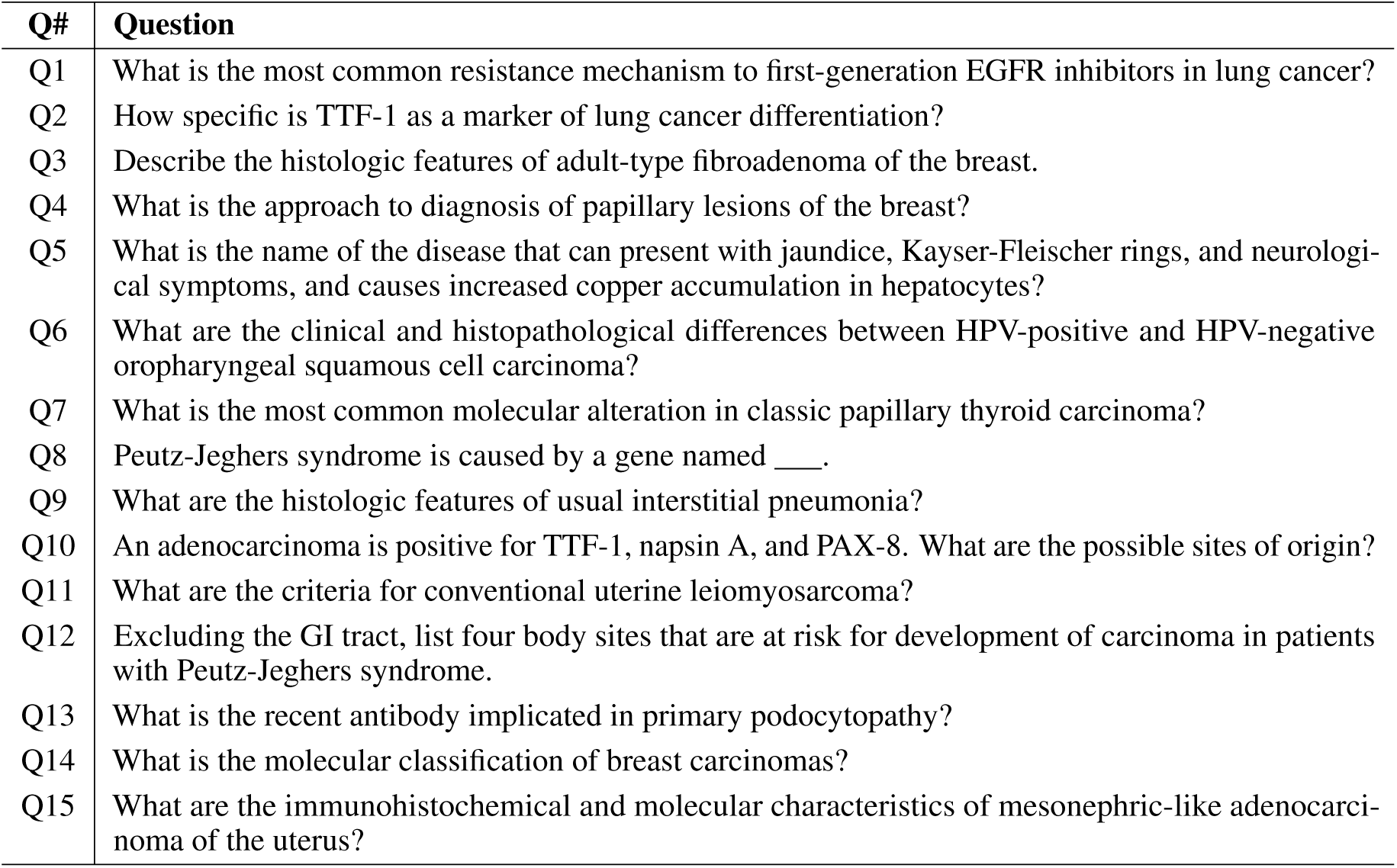
Diagnostic pathology questions used in the evaluation.

| Q# | Question |
| --- | --- |
| Q1 | What is the most common resistance mechanism to first-generation EGFR inhibitors in lung cancer? |
| Q2 | How specific is TTF-1 as a marker of lung cancer differentiation? |
| Q3 | Describe the histologic features of adult-type fibroadenoma of the breast. |
| Q4 | What is the approach to diagnosis of papillary lesions of the breast? |
| Q5 | What is the name of the disease that can present with jaundice, Kayser-Fleischer rings, and neurological symptoms, and causes increased copper accumulation in hepatocytes? |
| Q6 | What are the clinical and histopathological differences between HPV-positive and HPV-negative oropharyngeal squamous cell carcinoma? |
| Q7 | What is the most common molecular alteration in classic papillary thyroid carcinoma? |
| Q8 | Peutz-Jeghers syndrome is caused by a gene named ____. |
| Q9 | What are the histologic features of usual interstitial pneumonia? |
| Q10 | An adenocarcinoma is positive for TTF-1, napsin A, and PAX-8. What are the possible sites of origin? |
| Q11 | What are the criteria for conventional uterine leiomyosarcoma? |
| Q12 | Excluding the GI tract, list four body sites that are at risk for development of carcinoma in patients with Peutz-Jeghers syndrome. |
| Q13 | What is the recent antibody implicated in primary podocytopathy? |
| Q14 | What is the molecular classification of breast carcinomas? |
| Q15 | What are the immunohistochemical and molecular characteristics of mesonephric-like adenocarcinoma of the uterus? |

## Supplementary 3. Qualifications of Pathologists

**Table 2:**
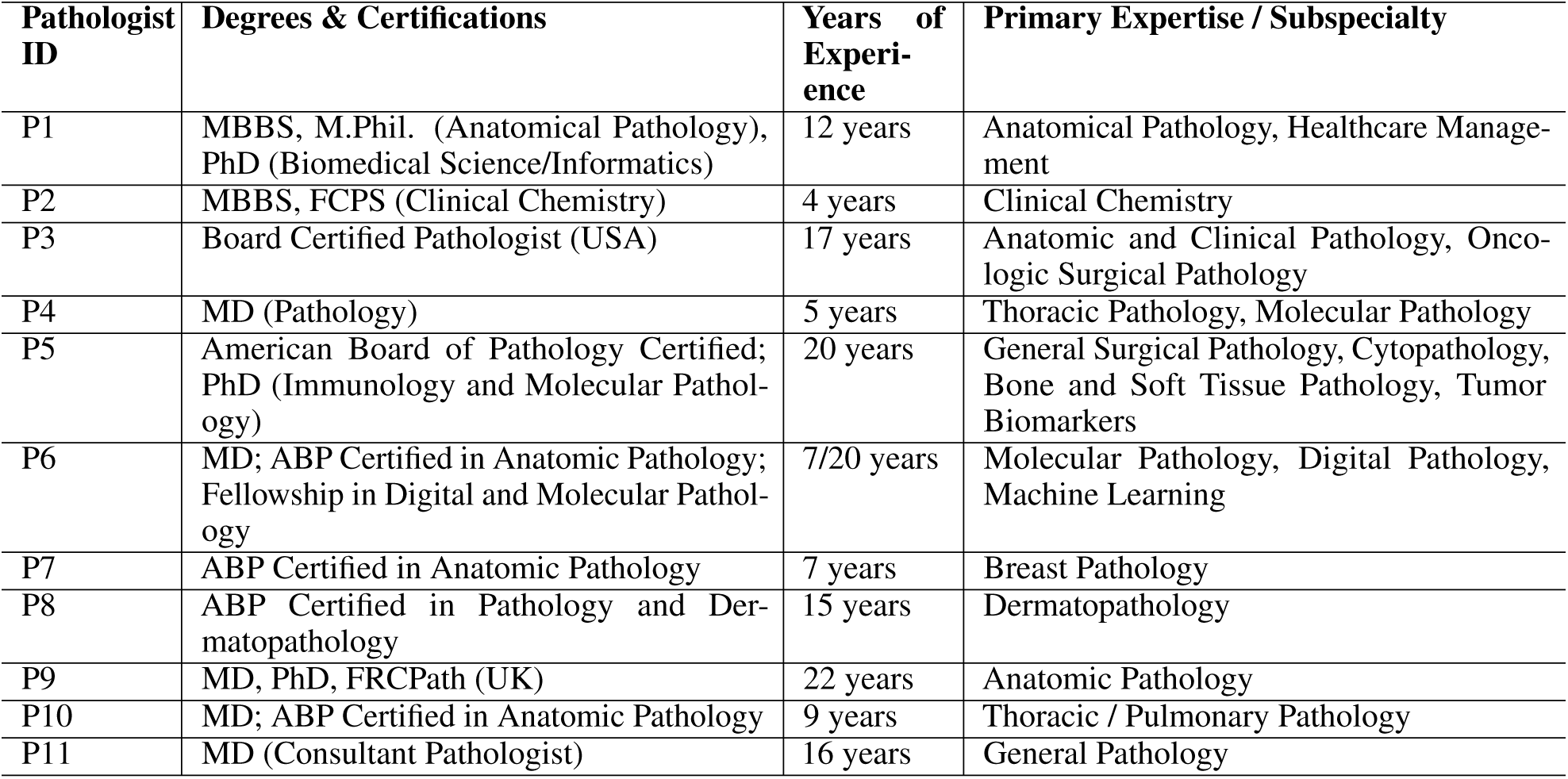
Summary of Pathologist Qualifications, Experience, and Subspecialties.

| Pathologist ID | Degrees & Certifications | Years of Experience | Primary Expertise / Subspecialty |
| --- | --- | --- | --- |
| P1 | MBBS, M.Phil. (Anatomical Pathology), PhD (Biomedical Science/Informatics) | 12 years | Anatomical Pathology, Healthcare Management |
| P2 | MBBS, FCPS (Clinical Chemistry) | 4 years | Clinical Chemistry |
| P3 | Board Certified Pathologist (USA) | 17 years | Anatomic and Clinical Pathology, Oncologic Surgical Pathology |
| P4 | MD (Pathology) | 5 years | Thoracic Pathology, Molecular Pathology |
| P5 | American Board of Pathology Certified; PhD (Immunology and Molecular Pathology) | 20 years | General Surgical Pathology, Cytopathology, Bone and Soft Tissue Pathology, Tumor Biomarkers |
| P6 | MD; ABP Certified in Anatomic Pathology; Fellowship in Digital and Molecular Pathology | 7/20 years | Molecular Pathology, Digital Pathology, Machine Learning |
| P7 | ABP Certified in Anatomic Pathology | 7 years | Breast Pathology |
| P8 | ABP Certified in Pathology and Dermatopathology | 15 years | Dermatopathology |
| P9 | MD, PhD, FRCPath (UK) | 22 years | Anatomic Pathology |
| P10 | MD; ABP Certified in Anatomic Pathology | 9 years | Thoracic / Pulmonary Pathology |
| P11 | MD (Consultant Pathologist) | 16 years | General Pathology |

## Supplementary 4. Definitions of Diagnostic Reasoning Strategies in Pathology

**Table 3:** Definitions of Diagnostic Reasoning Strategies in Pathology.

| Reasoning Strategy | Definition in the Context of Pathology |
| --- | --- |
| **Pattern Recognition** | The immediate identification of a diagnosis based on visual familiarity with histologic patterns. Common in routine diagnoses (e.g., basal cell carcinoma). Relies on accumulated experience and intuitive visual memory. |
| **Algorithmic Reasoning** | Stepwise application of decision trees or diagnostic algorithms. Each pathologic finding triggers a specific follow-up question or test, narrowing toward a final diagnosis (e.g., using immunostain panels for tumor typing). |
| **Inductive Hypothetico-Deductive Reasoning** | Formulating multiple diagnostic hypotheses based on initial findings and progressively narrowing them through targeted data gathering (e.g., special stains, clinical correlation). Involves both generating and testing diagnostic hypotheses. |
| **Bayesian/Probabilistic Reasoning** | Applying probabilistic thinking to estimate diagnostic likelihoods based on observed findings and known disease prevalence. Incorporates clinical evidence, test sensitivity/specificity, and prior probability. |
| **Deductive and Inductive Reasoning** | Deductive: applying general principles or rules to a specific case (e.g., if all tumors of type X express marker Y, a tumor that expresses Y may be type X). Inductive: building generalizations from specific observations (e.g., recognizing a new tumor variant). |
| **Heuristic Reasoning** | Use of mental shortcuts or experience-based rules to quickly arrive at likely diagnoses. While efficient, this approach may bypass systematic evaluation and can introduce bias. Appropriate caution is needed in modern diagnostic workflows. |
| **Mechanistic Insights** | Reasoning based on the underlying biological, molecular, or pathophysiological mechanisms of disease (e.g., linking KRAS mutation to mucinous colorectal adenocarcinoma phenotype). Enhances interpretation of complex or ambiguous findings. |

## Supplementary 5. Model Performance Across All Questions

This section presents per-question performance scores for each LLM, visualized separately for (1) language quality and structure and (2) diagnostic reasoning strategies. Each point represents the average normalized score from expert ratings across the four models on a single diagnostic question.

**Figure 6:**
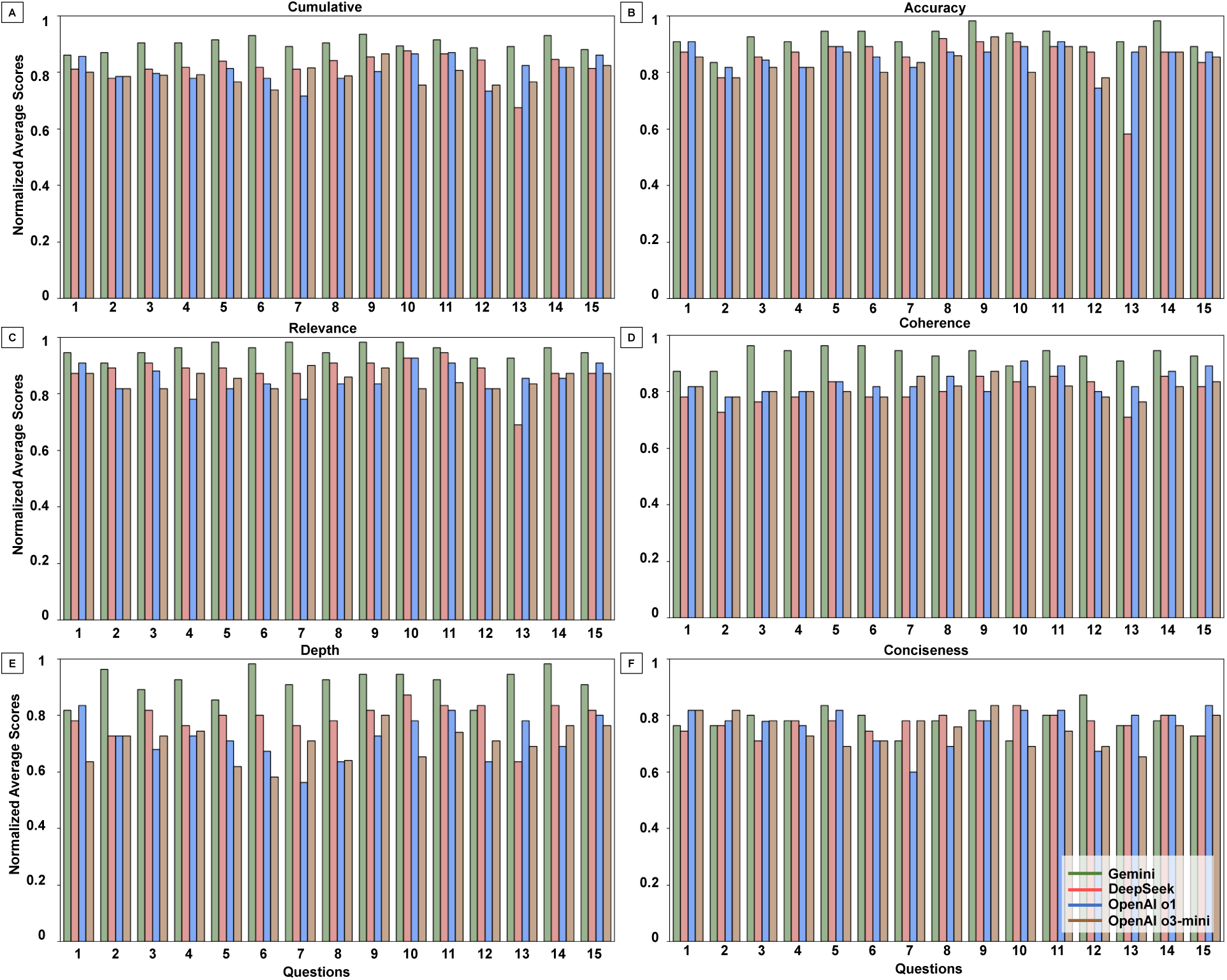
Pathologist Ratings of LLMs Across Language Quality Metrics. Normalized average scores (scale: 0–1) assigned by 11 pathologists across five natural language quality metrics: Accuracy, Relevance, Coherence, Conciseness, and Depth. Each point represents the mean score for a specific model on a single question. Gemini shows consistently strong performance, while other models exhibit greater variability across questions and metrics.

**Figure 7:**
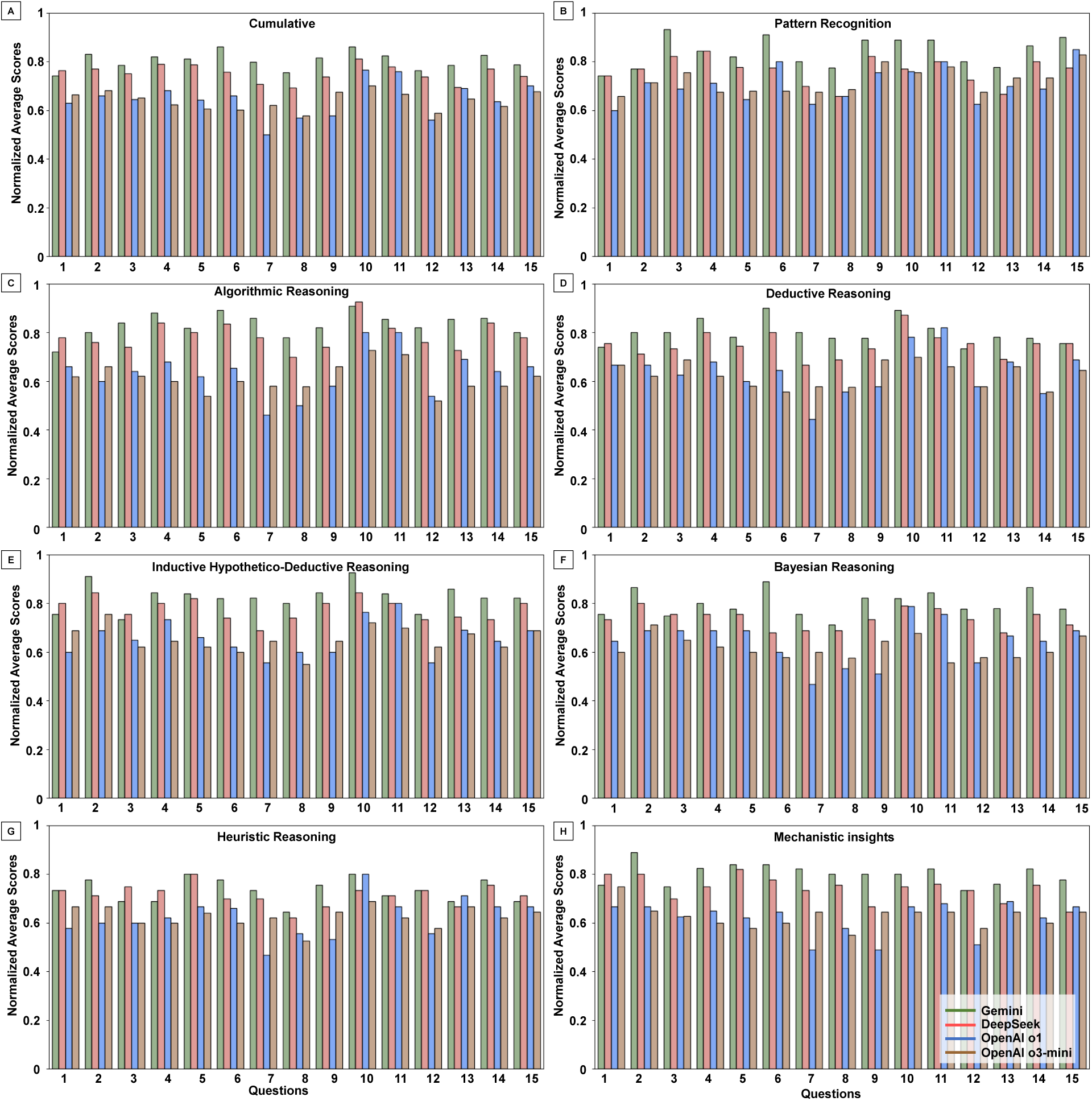
Pathologist Ratings of LLMs Across Diagnostic Reasoning Strategies. Normalized average scores (scale: 0–1) assigned across seven reasoning strategy criteria: Algorithmic, Pattern Recognition, Deductive, Inductive, Bayesian, Heuristic, and Mechanistic. Models vary in their use of domain-specific reasoning, with Gemini achieving higher and more consistent ratings across questions.

## Supplementary 6. Tukey’s Honestly Significant Difference (HSD) Pairwise Comparisons

**Table 4:** Pairwise Comparisons for Language Quality and Response Structure metrics.

|  | Metric | Comparison | p-value | Significant |
| --- | --- | --- | --- | --- |
| 1 | Relevance | Gemini vs DeepSeek | < 0.0001 | Yes |
| 2 | Relevance | Gemini vs OpenAI o1 | < 0.0001 | Yes |
| 3 | Relevance | Gemini vs OpenAI o3-mini | < 0.0001 | Yes |
| 4 | Relevance | DeepSeek vs OpenAI o1 | 0.2054 | No |
| 5 | Relevance | DeepSeek vs OpenAI o3-mini | 0.1935 | No |
| 6 | Relevance | OpenAI o1 vs OpenAI o3-mini | > 0.9999 | No |
| 7 | Coherence | Gemini vs DeepSeek | < 0.0001 | Yes |
| 8 | Coherence | Gemini vs OpenAI o1 | < 0.0001 | Yes |
| 9 | Coherence | Gemini vs OpenAI o3-mini | < 0.0001 | Yes |
| 10 | Coherence | DeepSeek vs OpenAI o1 | 0.0847 | No |
| 11 | Coherence | DeepSeek vs OpenAI o3-mini | 0.8825 | No |
| 12 | Coherence | OpenAI o1 vs OpenAI o3-mini | 0.3408 | No |
| 13 | Depth | Gemini vs DeepSeek | < 0.0001 | Yes |
| 14 | Depth | Gemini vs OpenAI o1 | < 0.0001 | Yes |
| 15 | Depth | Gemini vs OpenAI o3-mini | < 0.0001 | Yes |
| 16 | Depth | DeepSeek vs OpenAI o1 | 0.0108 | Yes |
| 17 | Depth | DeepSeek vs OpenAI o3-mini | 0.0009 | Yes |
| 18 | Depth | OpenAI o1 vs OpenAI o3-mini | 0.8415 | No |
| 19 | Accuracy | Gemini vs DeepSeek | 0.0046 | Yes |
| 20 | Accuracy | Gemini vs OpenAI o1 | 0.0076 | Yes |
| 21 | Accuracy | Gemini vs OpenAI o3-mini | 0.001 | Yes |
| 22 | Accuracy | DeepSeek vs OpenAI o1 | 0.9982 | No |
| 23 | Accuracy | DeepSeek vs OpenAI o3-mini | 0.9579 | No |
| 24 | Accuracy | OpenAI o1 vs OpenAI o3-mini | 0.9057 | No |
| 25 | Conciseness | Gemini vs DeepSeek | 0.9804 | No |
| 26 | Conciseness | Gemini vs OpenAI o1 | 0.8648 | No |
| 27 | Conciseness | Gemini vs OpenAI o3-mini | 0.4136 | No |
| 28 | Conciseness | DeepSeek vs OpenAI o1 | 0.9794 | No |
| 29 | Conciseness | DeepSeek vs OpenAI o3-mini | 0.6494 | No |
| 30 | Conciseness | OpenAI o1 vs OpenAI o3-mini | 0.8648 | No |
| 31 | Cumulative | Gemini vs DeepSeek | 0.0212 | Yes |
| 32 | Cumulative | Gemini vs OpenAI o1 | < 0.0001 | Yes |
| 33 | Cumulative | Gemini vs OpenAI o3-mini | < 0.0001 | Yes |
| 34 | Cumulative | DeepSeek vs OpenAI o1 | < 0.0001 | Yes |
| 35 | Cumulative | DeepSeek vs OpenAI o3-mini | < 0.0001 | Yes |
| 36 | Cumulative | OpenAI o1 vs OpenAI o3-mini | 0.9901 | No |

**Table 5:**
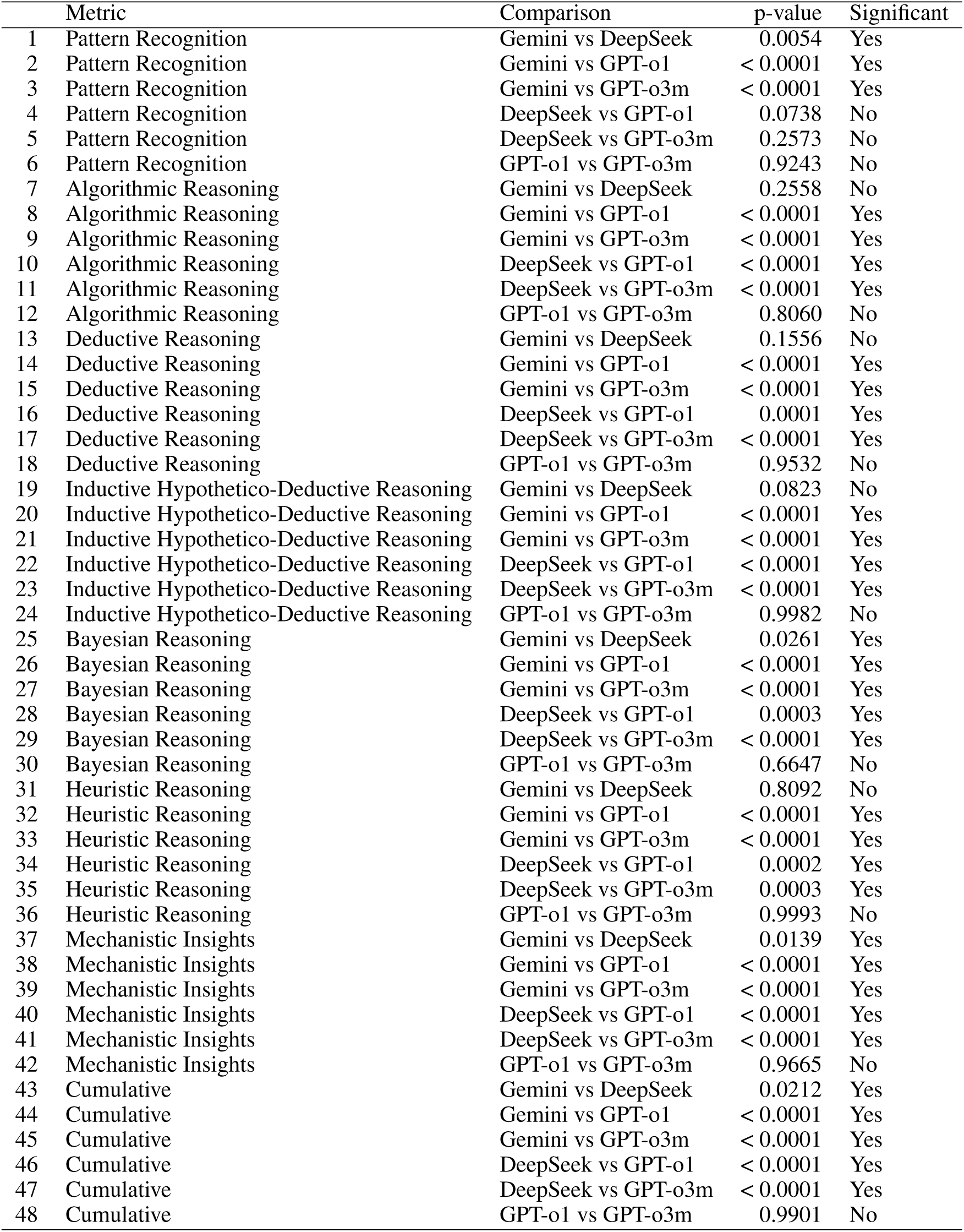
Pairwise Comparisons for Diagnostic Reasoning Strategies.

## Supplementary 7. Inter-observer Agreement Analysis

**Table 6:** Summary of Percent Agreement Across Models. Question-Model-Criteria (Q-M-C)

| **Model** | **Mean Percent Agreement (SD)** | **Q-M-C with Percent Agreement < 0.4** |
| --- | --- | --- |
| Gemini | 0.65 (0.12) | 15 |
| DeepSeek | 0.43 (0.09) | 37 |
| OpenAI o1 | 0.42 (0.10) | 38 |
| OpenAI o3-mini | 0.41 (0.08) | 29 |

## Supplementary 8. Missing Score Analysis and Normalization Approach

To assess data completeness, we quantified the number of missing evaluations per pathologist across all question–model–criterion (Q–M–C) combinations. The distribution of missing scores was highly skewed, with the majority originating from two evaluators: Pathologist-10 (341 missing entries) and Pathologist-8 (339). All other evaluators contributed near-complete data, with fewer than 35 missing values each. Seven of the eleven evaluators had fewer than 20 missing scores, and three (Pathologists 3, 9, and 11) each had only a single missing entry.

To ensure fair comparison across models, we normalized scores for each Q–M–C combination by computing the mean of available ratings only. This approach prevents penalizing models for missing evaluations and ensures that aggregate scores reflect only completed assessments. Importantly, all Q–M–C combinations had a minimum of seven independent evaluations, with a mean of 9.9 ratings per combination, supporting the robustness of model-level comparisons despite occasional missing data.

**Table 7:** Missing score counts and percentages by model.

| Model | Total Scores | Missing Count | Missing % |
| --- | --- | --- | --- |
| Gemini | 1980 | 177 | 8.94% |
| DeepSeek | 1980 | 179 | 9.04% |
| OpenAI o1 | 1980 | 198 | 10.00% |
| OpenAI o3-mini | 1980 | 218 | 11.01% |
| **Total** | **7920** | **772** | **9.75%** |

**Table 8:** Missing score counts and percentages by question.

| Question | Total Scores | Missing Count | Missing % |
| --- | --- | --- | --- |
| Q1 | 528 | 61 | 11.55 |
| Q2 | 528 | 61 | 11.55 |
| Q3 | 528 | 70 | 13.26 |
| Q4 | 528 | 54 | 10.23 |
| Q5 | 528 | 36 | 6.82 |
| Q6 | 528 | 39 | 7.39 |
| Q7 | 528 | 56 | 10.61 |
| Q8 | 528 | 71 | 13.45 |
| Q9 | 528 | 52 | 9.85 |
| Q10 | 528 | 32 | 6.06 |
| Q11 | 528 | 37 | 7.01 |
| Q12 | 528 | 56 | 10.61 |
| Q13 | 528 | 36 | 6.82 |
| Q14 | 528 | 53 | 10.04 |
| Q15 | 528 | 58 | 10.98 |

**Table 9:** Missing score counts and percentages by evaluation criterion.

| Criterion | Total Entries | Missing Count | Missing % |
| --- | --- | --- | --- |
| Clarity | 660 | 4 | 0.61 |
| Coherence | 660 | 2 | 0.30 |
| Depth | 660 | 3 | 0.45 |
| Accuracy | 660 | 5 | 0.76 |
| Conciseness | 660 | 3 | 0.45 |
| Pattern Recognition | 660 | 151 | 22.88 |
| Algorithmic Reasoning | 660 | 46 | 6.97 |
| Inductive Reasoning | 660 | 94 | 14.24 |
| Mechanistic Reasoning | 660 | 103 | 15.61 |
| Deductive Reasoning | 660 | 117 | 17.73 |
| Heuristic Reasoning | 660 | 120 | 18.18 |
| Probabilistic Reasoning | 660 | 124 | 18.79 |

## Notes

### Competing Interest Statement

The authors have declared no competing interest.

### Summary of Updates

Revised Figs 3 and 4. Extended Discussion section. Added two new sections to the Supplementary material.

